# Double-blind, Randomized, Placebo-Controlled study to evaluate the Efficacy of an early treatment with Herbal Supplement based in the Prevention of Post-Traumatic Stress Disorder in emergency department (PHYTéS Study)

**DOI:** 10.1101/2022.01.12.22268879

**Authors:** Riadh Boukef, Rym Youssef, Hajer Yaakoubi, Imen Trabelsi, Adel Sekma, Khaoula Bel Hadj Ali, Houda Ben Salah, Amal Baccari, Lotfi Boukadida, Asma Zorgati

## Abstract

**Introduction:** The prevention from Post-traumatic stress disorder (PTSD) is therefore of major public health interest and one of the concerns of any emergency physician. The purpose of our study was to evaluate the efficacy and safety of an herbal supplement to prevent the occurrence of PTSD in high-risk patients.

**Methods:** It is a randomized, double-blind, prospective, interventional study including patients exposed to a potentially traumatic event that meets DSM-V Criterion A and has a Peri-traumatic Distress Inventory score or the Questionnary for traumatic dissociation experiments (PDEQ) and/or L.Crocq score higher than the thresholds between day 1 and day 3. Two hundred patients were included randomly assigned into two groups: Aleozen group and placebo group. Patients included in aleozen group received Aleozen^®^ for 10 days while patients in placebo group received Placebo. A CAPS-5 assessment was performed for all patients at different moments. The main objective was to assess the efficacy of Aleozen^®^ after 90 days of an exposition to traumatic events according to PTSD. Secondary objectives were to evaluate the safety of Aleozen^®^ at 10 and 30 days after its administration and PTSD in the study population after one year of inclusion.

**Results:** No statistical differences were noted between the two groups in term of baseline characteristics including age, sex and the ISS score. After 90 days of follow-up, and according to the CAPS-5 scale, 85 patients (42.5%) of the population study showed PTSD. Concerning primary endpoint, less PTSD were seen in intervention group compared to placebo group (38.8% versus 61.2% respectively; p<0.001). During the study, no adverse events were noted.

**Conclusion:** Results of this work suggest the potential preventive effects of an herbal supplement on PTSD for traumatic patient in emergency. Further confirmatory studies are needed.

**Funding sources:** This research is supported by « Aleonat » Laboratories, who provided only the protocol treatments (active and placebo), without other financial support of the study. The sponsor and funder have no role in study design; collection, management, analysis, and interpretation of data; writing of the report; and the decision to submit the report for publication, including whether they will have ultimate authority over any of these activities.

**Ethical approval:** The study was approved by the local ethics committee (The Ethics Committee of the Faculty of Medicine of Sousse)

**Ethical review:** A written informed consent was obtained from all participants.

**Human and animal rights statement:** All procedures performed in the study were in accordance with the ethical standards of the institutional and/or national research committee and with the 1964 Helsinki Declaration and its later amendments or comparable ethical standards.

## Introduction

Trauma-related nightmares and sleep disturbance are common symptoms of post-traumatic stress disorder (PTSD) [1]. It’s a debilitating psychopathological response to a traumatic event which can severely compromise quality of life. In recent years, the increasing of estimated prevalence of PTSD can be due partly to the improvement of the standardized evaluation procedure.[1]

One of the first epidemiological studies of PTSD was “Mental health in general population” survey which was carried in metropolitan France between 1999 and 2003 including 36,000 people. This study revealed that PTSD instantaneous prevalence (last month) was 0.7% of the population with nearly equal average between men (45%) and women (55%) [2]. Same result was reported in the European study of the epidemiology of mental disorders [3]. Moreover, several psychiatric and physical comorbidities may occur with PTSD, particularly mood disorders, anxiety, and substance abuse [4]. Nevertheless, its severity is worsened by the high risk of suicidal behavior [5]. Moreover, one of the first published studies that assessed the relation between exposure to war, PTSD and mortality was conducted among Vietnam veterans in the United State. Veterans with elevated level of PTSD risk have a highest mortality risk (adjusted hazard ratio = 2.34, 95% confidence interval: 1.24, 4.43) [6]. Definition of PTSD was conflictual; Barrois defines this traumatic neurosis as “a group of psychological disorders that arise after a longer or shorter latency, following a very intense emotional shock” [7].

Anxiolytic and/or sedative phytotherapy may represent an alternative symptomatic treatment for anxiety states which are not too severe. Thus, in its recommendations for out-patient management of anxiety disorders in adults, the ANAES (Agence Nationale d’Accréditation et d’Evaluation en Santé) supports the use of a phytotherapy preparation, Euphytose, based on dry extracts of Crataegus, Balloia, Passiflora, Valeriana, Cola and Paullinia [8].

The burden of several diseases associated with psycho traumatic disorders highlight the importance of prevention and primary care which remains under-identified, particularly in Tunisia. So, several therapeutic protocols have been proposed over the years. The antidepressant treatment (yet first-line treatment in all international guidelines) represents only 30% of PTSD treatment and the remaining 70% include anxiolytics, hypnotics and herbal medicine. Several meta-analyzes have evaluated the efficacy of antidepressants, but data for herbal medicine remains very poor. We tested Aleozen^®^, a balanced and synergistic combination of plant extracts growing in Tunisia that helps treat states of stress, insomnia, and anxiety [9-11].

This study aims to evaluate the efficacy and the tolerance of Aleozen^®^) versus placebo in patients with high risk of developing PTSD.

## Methods

A double-blind, randomized placebo-controlled clinical trial was conducted from June 2018 to September 2018. The study was approved by the Ethics Committee; it was recorded in the ClinicalTrials.gov register.

The treatment was provided by Aleonat Laboratories, Tunisia.

### Population

Patients, over 18 years of age, who had been exposed to a traumatic event (a violent mechanism such as assault, public road or domestic accident) meeting criterion A of the definition of PTSD according to DSM-V [10] were included.

Included patients had a total score of Peri-traumatic Distress Inventory (PDI)score>15 [12] and/or the Peri-traumatic Dissociative Experiences Questionnaire (PDEQ)>15 [13] and/or the Immediate Stress Questionnaire (L. Crocq score) [12]>50 between Day1 and Day3 after the traumatic event.

PDI is a 13-item self-report scale for determining a person’s emotional distress reactions, at the time of a traumatic event and in the minutes and hours after it. People with severe peri-traumatic distress are at risk of developing post-traumatic stress disorder. A score greater than 15 indicates significant dissociation [12].

The PDEQ has been validated in French and uses the English version. It is a self-administered questionnaire to retrospectively assess the dissociative elements of consciousness at the time of the trauma and in the following minutes in 10 items. A score greater than 15 indicates significant dissociation [13].

L. Crocq is a self-administered questionnaire of 20 questions, exploring the five aspects cognitive, affective and mental state immediately after the event; A score greater than 50 indicates significant dissociation [14].

Individuals with serious trauma admitted to the intensive care unit, patients with psychiatric illness or using psychotropic medication, non-cooperating patient and non-consenting patients were not included.

About the sample size to estimate the minimum sample size, we used the following formula n = [3.84 * ((P * Q) / i2)] with P = the prevalence of PTSD = 0.15, Q = 1-P = 0.85, i = 0.05, or 195 patients.

We supposed that the prevalence of PTSD was about 15% according to the study of Van Zuiden et al where the prevalence of the PTSD was found= 18,44% and in a second reason to a study carried out in our department (not yet published) including patients with the same criteria as our current study (patient at high risk of manifesting PTSD) the prevalence was found to be 14% [15].

### Study design

Participants were randomized to 10 days double-blind treatment with ALEOZEN^®^ or placebo. Aleozen^®^ capsula contains *Griffonia simplicifolia* and *Rhodiola rosea* with a dosage of 110 mg per capsule each. Second, *Gentiana lutea* with a dose of 100 mg per capsule. *Crataegus oxyacantha, Eschscholtzia californica* and *Melissa officinalis* are present at a dose of 60 mg each. The capsule also contains Zinc Bisglycinate and vitamin B6. The ALEOZEN^®^ and placebo treated patients received 1 capsule*3 times per day. ALEOZEN^®^ and placebo capsule were identical shape and color. Randomization was performed according to a 4:4 random survey.

### Data collection

Data was collected by medical staff practicing at Sahloul emergency department. On the first day of admission to the emergency room, a pre-tested questionnaire with three items was used for data collection.

The first item treated demographic characteristics such as age, sex or medical history.

The second one included the elements of physical examination; traumatic lesions and the injury severity score (ISS) [16]. In the third part we tried to assess the severity of the traumatic event using the PDEQ [13], the PDI scale [12] and the Immediate stress questionnaire (L. Crocq) [14].

The follow-up of the patient was carried on the 10th day, 1month, 3month and one year after the study inclusion via a telephone consultation. These calls were made by a researcher who was blind to the treatment taken. Double-blind anonymity was issued after statistical analysis of all data. The main objective was to confirm the diagnosis of PTSD according to the Clinically Administered PTSD Scale for DSM-5 (CAPS-5 scale) [17] at the third month. At the 10^th^ day, telephone call was made ensuring the safety and the adherence to the treatment; at one month, we wanted to evaluate its safety and effectiveness. At 3 months, the calls were made to evaluate PTSD. At one year after inclusion to the study, the purpose of the telephone call is to evaluate the progress of patients admitted to each group and assess the progress of patients according to their post-traumatic stress disorder (figure 2). We adopted score CAPS-5 >32 to retain PTSD diagnosis [18]. CAPS-5 is a self-administered 20-item questionnaire that evaluates the 20 symptoms of PTSD according to the CAPS-5. A provisional diagnosis can be obtained by considering any item with a score of 2 or more as present, and then adhering to CAPS-5 instructions that require at least: 1 item B (questions 1-5), 1 item C (questions 6-7), 2 items D (questions 8-14), and 2 items E (questions 15-20). A threshold of 38 at CAPS-5 seems reasonable to suggest the presence of PTSD until psychometric studies are available.

**Figure 1:**
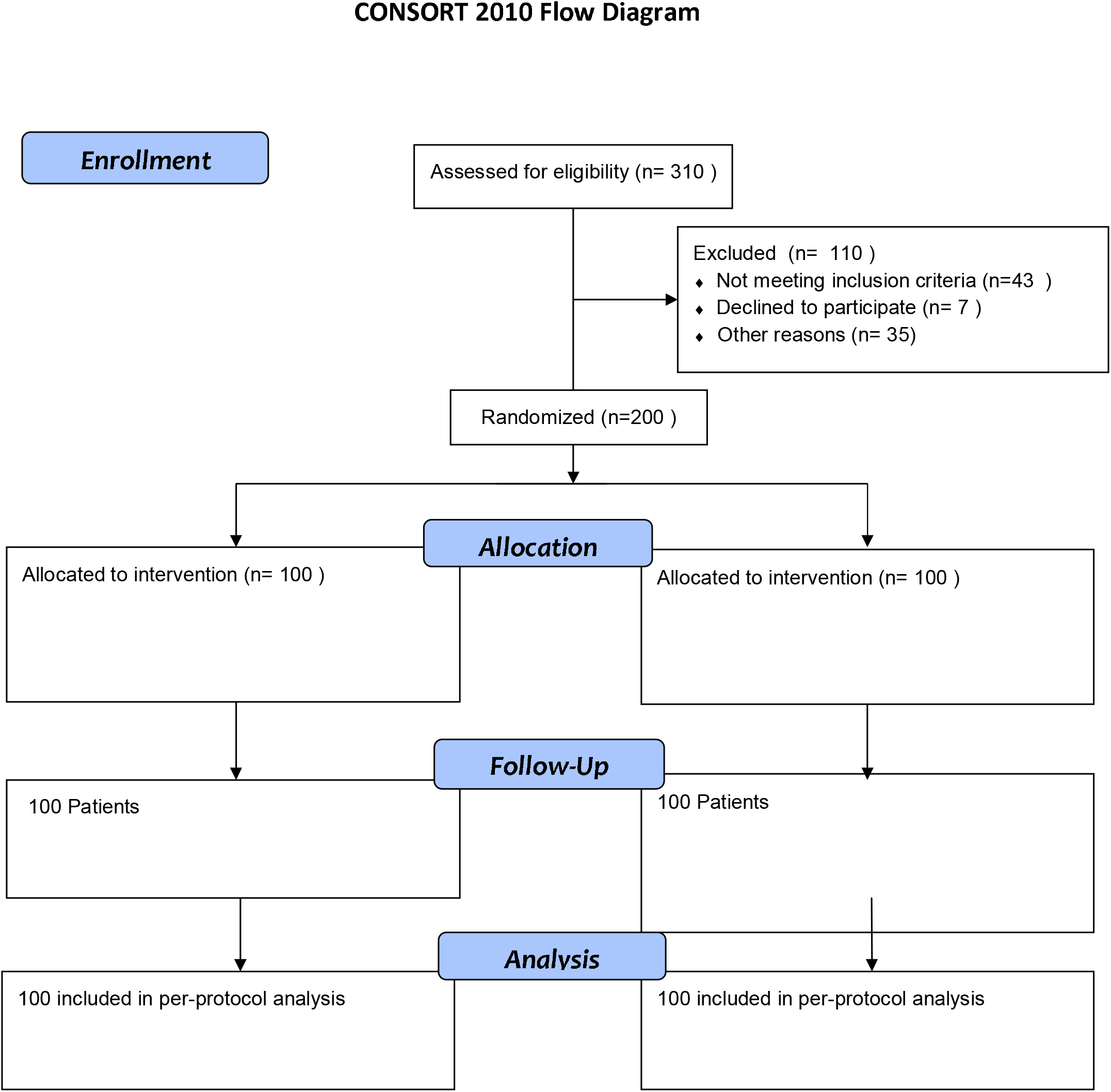
CONSORT 2010 Flow Diagram.

**Figure 2:**
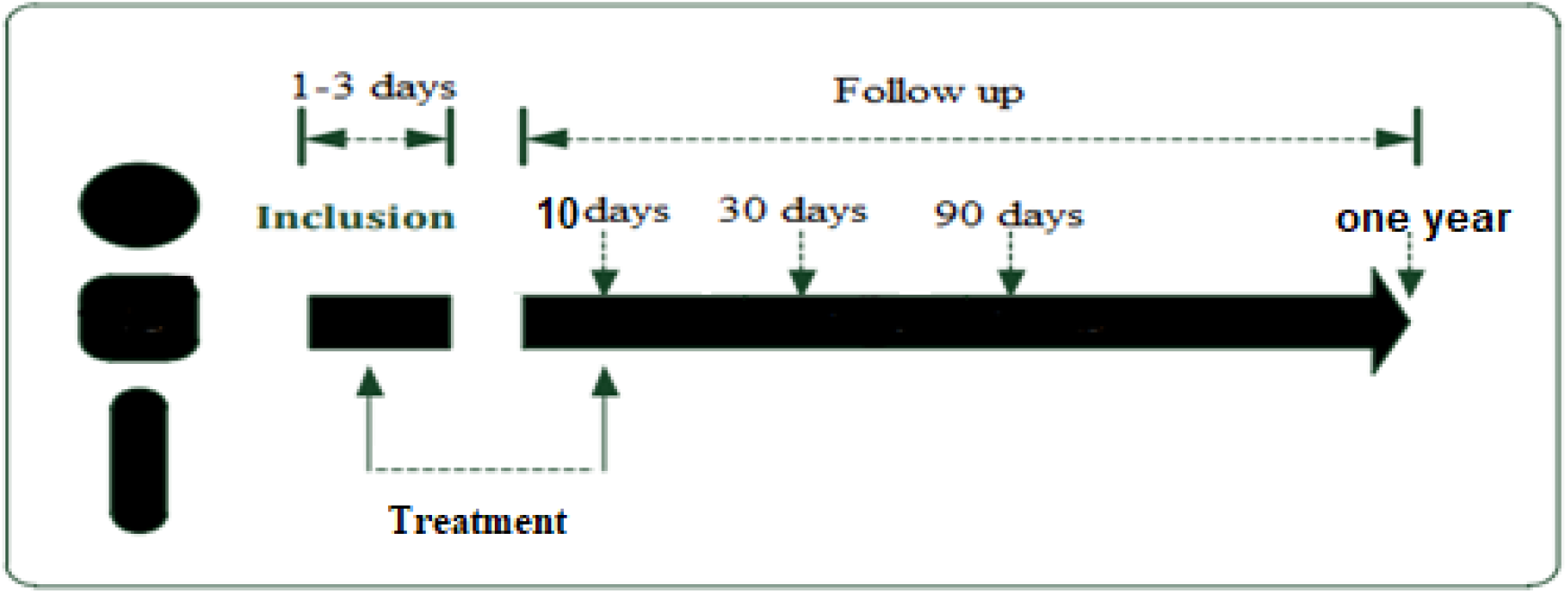
Study protocol.

### Endpoints of the study

The primary endpoint was to assess improvement of PTSD at 90 days after the beginning of Aleozen therapy. The secondary objectives were to evaluate the safety of Aleozen^®^ at 10 and 30 days after its administration and efficacy of PTSD after one year of inclusion. We evaluated also the improvement of symptoms severity of PTSD (reduction of the CAPS-5 score by 50%), and necessity of psychiatric follow-up, use of additional psychotropic or anxiolytic treatment or adverse events.

### Statistical analysis

The data was entered and analyzed using the Statistical Package for Social Sciences (SPSS). Categorical variables were reported as count and percentages. Quantitative variables were expressed in terms of means and standard deviation. For the comparison of the qualitative variables, we used the Chi2 test and for the comparison of the quantitative variables, Student’s and ANOVA test. Variables having p-value of less than 0.05 were considered as significant at 95% confidence level.

## Results

We included 200 patients in our study: 100 patients in each group. The characteristics of participants are shown in table1. No statistical differences were noted between the two groups in term of age, sex and the ISS score. Overall, 8.5% of the participants were lost to follow-up. After 90 days of follow up, and according to the CAPS-5 Scale, 85 patients (42,5%) of all the study population presented PTSD. Concerning primary endpoint, less PTSD were seen in Aleozen^®^ group compared to placebo group (38.8% versus 61.2% respectively, p<0.001). Patients with PTSD were referred either to psychiatrists or to a psychologist working in the emergency rooms according to the patient’s choice.

**Table1:**
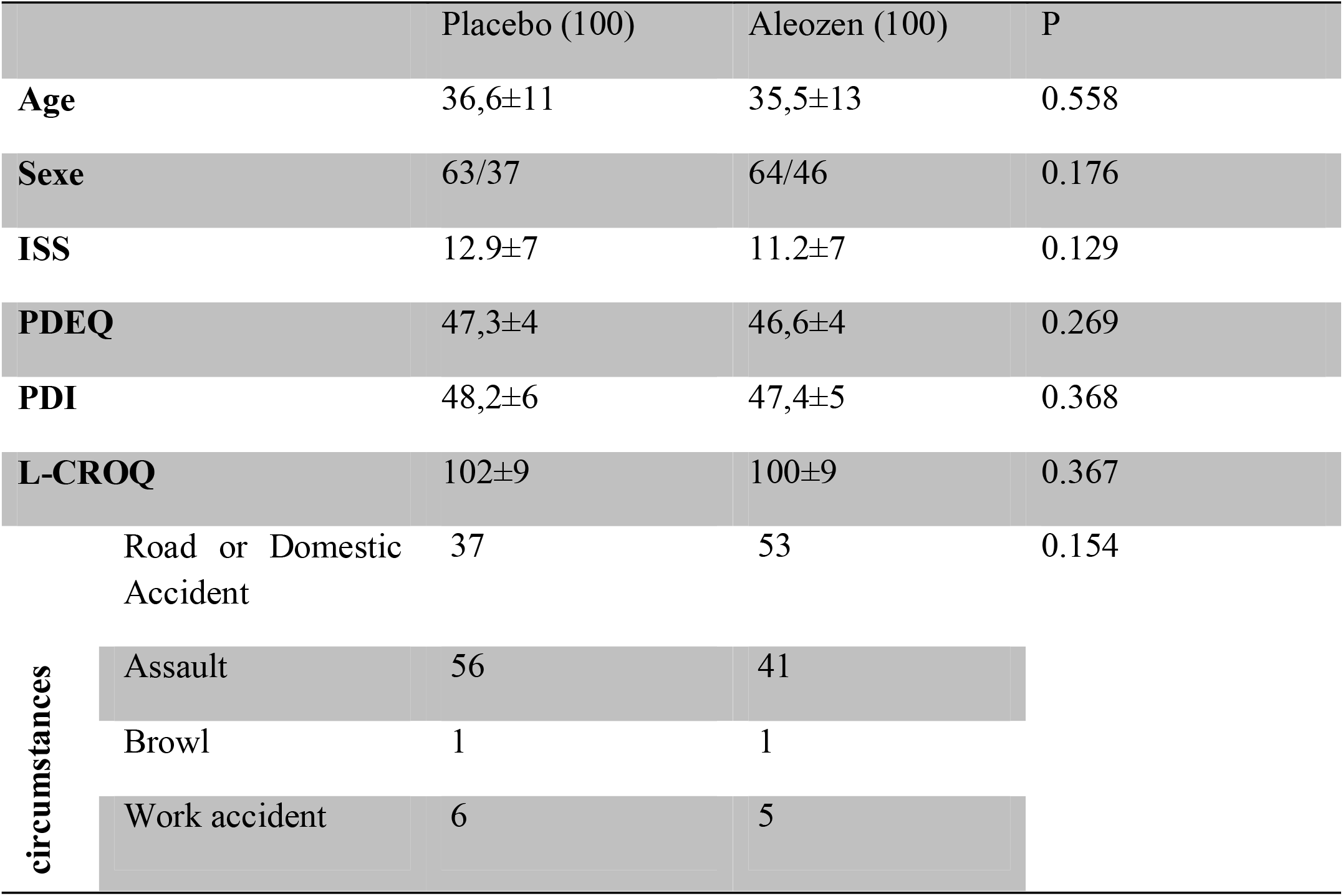
Population’s characteristics.

A decrease in the total rate of CAPS-5 by 50% was observed mostly among patients in the Aleozen^®^ group (51% VS 29%, p=0.004).

When comparing the CAPS-5 scale between the two groups; no significant differences were noted 10 and 30 days after inclusion. However, a significant difference appeared in day 90 (40.6±2 for aleozen group vs 54.7±24 for placebo group, p<0.001) and after one year (38.6±12 versus 47.8±14 respectively, p=0.032) (figure 3). In total, 88 patients had a decrease of at least 50% in CAPS-5 at 3 months; 59 (67%) patients of Aleozen group versus 29 (33%) patients of placebo group (p<0. 05).

**Figure 3:**
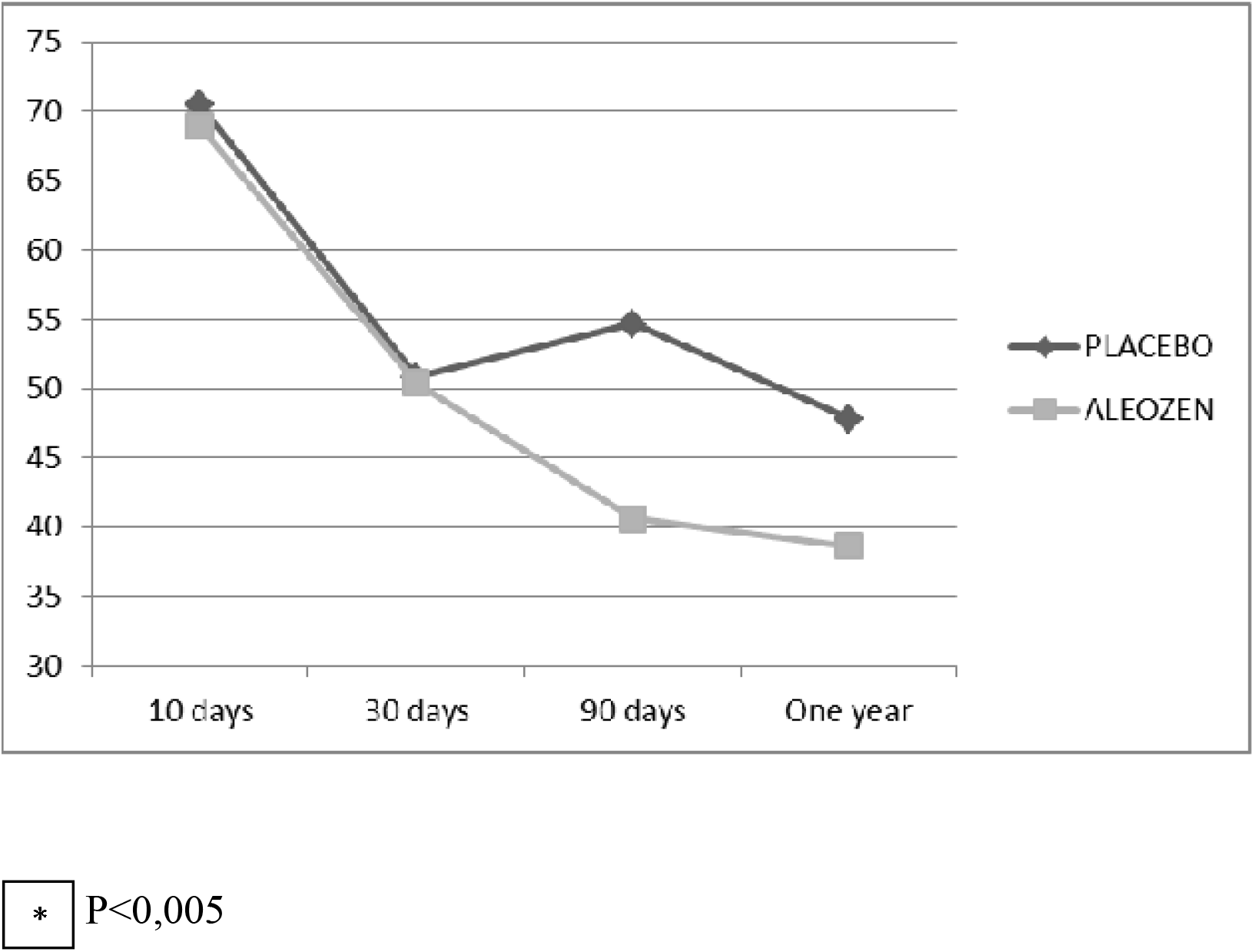
Evolution of the mean of CAPS-5 between Aleozen and Placebo groups over time.

The comparison between the 4 items of CAPS 5 showed almost similar results between the two groups after 10 days and 30 days of inclusion. However, a significant difference was found after 90 days and one year of follow up, particularly between B, C and D items.

When observing the triggering mechanism of ED admission, a significant benefit of Aleozen® was noted in traffic accident and assault among the two groups (p=0.025 and 0.006 respectively).

The variation of CAPS-5 in relation to the admission has been shown in table 2. Observation a comparable variation at 30 days against two significant differences in the variation of the CAPS at 90 days and one year

**Table 2:** CAPS-5 variations.

| **Table 2 : CAPS-5 variations** |  |  |  |  |
| --- | --- | --- | --- | --- |
|  | Groups | Moyenne | SD | P |
| CAPS10days<br>(200 patients) | PLACEBO | 70.5 | 19.9 | 0.591 |
|  | ALEOZEN | 68.9 | 20.2 |  |
| CAPS30 days<br>(200 patients) | PLACEBO | 50.8 | 19.7 | 0.897 |
|  | ALEOZEN | 50.4 | 21.4 |  |
| CAPS 90 days<br>(191 patients) | PLACEBO | 54.7 | 24.4 | **<0.001** |
|  | ALEOZEN | 40.6 | 20.6 |  |
| CAPS One year<br>(183 patients) | PLACEBO | 47.8 | 14 | **0.032** |
|  | ALEOZEN | 38.6 | 12 |  |
| VARIATION 30 days | PLACEBO | 20.2 | 19.9 | 0.568 |
|  | ALEOZEN | 18.5 | 20.6 |  |
| VARIATION 90 days | PLACEBO | 2.7 | 30.1 | **0.001** |
|  | ALEOZEN | -10.4 | 21.8 |  |
| VARIATION one year | PLACEBO | 5.6 | 18.8 | **0.024** |
|  | ALEOZEN | -6.7 | 18.5 |  |

The comparison between the 4 items of CAPS5 showed almost similar results between the two groups of studies after 10 days, 30 days and one year of inclusion. However, after 90 days and one year of follow up, the differences were significant. (Table 3)

The difference between the 30- and 90-day CAPS-5 items, compared with the 10-day results, showed significant difference between the group of patients who received a placebo and the group who received the Aleozen^®^ always after 90 days of inclusion in items B, C and D (table 4).

After one year following up, and according to the CAPS-5 criteria, PTSD was diagnosed among 12% of placebo group and 6% from Aleozen^®^ group. These patients were referred to psychiatrists or psychologist. Five patients from placebo group were put on anxiolytics. During the study, no adverse events were noted.

## Discussion

In this randomized, double-blind, prospective study, our result strongly suggest benefits of a herbal medicine administration early after trauma on prevention of PTSD or reduction of subsequent PTSD symptom severity in emergency department in high-risk patients.

We showed in our study that prescribing Aleozen^®^ in emergency department for high risk patients for developing PTSD can prevent this stress disorder. For screening high risk patient we used 2 tools: PDI [12] and PDEQ scales. Those tools assessed peri-traumatic distress and peri-traumatic dissociation. While searching in the literature, we have found that they are the two main risk factors that were associated with development of PTSD. A meta-analysis by Ozer et al in 2003 which included 68 articles between 1980 and 2000 found that 16 studies have examined peri-traumatic dissociation as risk factors for PTSD. Of the 16 studies, the correlation between dissociative experiences during or immediately after traumatic exposure and symptoms of PTSD ranged from 0.14 to 0.94 with an adjusted mean correlation of 0.35 in the cumulative sample (n = 3534 subjects) [17]. Thus, distress and peri-traumatic dissociation measured by PDI [12] and PDEQ [13] tools respectively are statistically associated with the development of PTSD in adult populations. Indeed, we used in our study, Posttraumatic Check List 5 or CAPS-5 to assess the diagnosis of PTSD which is a revised scale of the CAPS DSM-IV [20]. Among several investigators who were interested in evaluating CAPS-5 scale, Wortman et al in 2016 [21], Marmar CR et al in 2015 [22] and Kruger Gottschalk et al in 2017 [23] showed that this scale is a strong psychometric tool for making a provisional diagnosis, to quantify the severity of PTSD symptoms and monitor clinical change over time witch make it the gold standard.Thus, it’s important to understand neurobiological mechanisms involved in generating PTSD to provide challenges for treatment. In the first studies in patients with PTSD, researchers showed autonomic reactivity in response to trauma-related signs, such as increased heart rate and skin conductance [24, 25]. Moreover, many pharmacological challenge trials with yohimbine (an α2-adrenergic receptor antagonist) demonstrated excessive neurochemical then behavioural reaction in relation with central noradrenergic hyper-reactivity in PTSD [26, 27]. As we understand that the pathophysiology of psycho-trauma involves several pathways, neurotransmitters and hormones, with clearly impact on nervous system and memory, these findings have been the subject of numerous studies. These are indeed promising avenues for developing pharmacological treatments to prevent the occurrence of post-traumatic stress disorder [28, 29]. Many pharmacological treatments are now available [30]. However, psychological approaches to treat PTSD still the gold standard and accordingly have been recommended as first line PTSD treatment by guidelines [31-33].

From this perspective, Zohar et al. conducted a small randomized trial to evaluate the efficacy of a single bolus of high-dose hydrocortisone (100 to 140 mg depending on weight) to prevent PTSD after mild trauma in patient who had symptoms of acute stress in the emergency department. Patients who had received hydrocortisone (n = 8–10) had significantly fewer PTSD symptoms than patients in the placebo group (n = 7–9) after 2 week and the first- and third-month follow-up [34].

More recently, Delahanty et al conducted a randomized controlled double-blind hydrocortisone vs placebo study for ten days, initiated twelve hours post-trauma in high risk PTSD patient. This study suggests that early treatment with low dose hydrocortisone low may be effective in preventing PTSD [35].

Besides, other studies were interested in B-blockers in particularly propranolol due to their role of interruption post-synaptic norepinephrine receptors. In this regard, Hoge et al conducted in 2012 a randomized, double-blind, emergency room study that included 41 patients: propranolol group and placebo group. However, propranolol did not show a superiority efficacy over placebo in PTSD [36]. Same results were found in a recent meta-analysis including 5 studies. However, the studies considered in this meta-analysis included samples of small numbers which could explain the absence of significant difference [37].

In addition, more studies have found beneficial effect for use of morphine [38], oxytocin [15, 29] and ketamine [39].

It has been attempting to compare different treatment approaches in PTSD. On this basis, a meta-analysis was conducted comprising 12 randomized clinical studies including 922 participants’ demonstrated similar results for the 3 approaches at the end of treatment. However long-term benefits of psychotherapeutic and combined treatments were superior to pharmacological treatments in 6 randomized clinical trials that reported follow-up data [40]. Otherwise, very few trials have investigated herbal medicine efficacy in prevention of PTSD and our study comes in this perspective. Numata and al in 2014 in Japan have conducted a controlled randomized double blind trial including 43 patients with an Impact Of Events Scale-Revised IES-R over 25. These patients have received either a herbal treatment, saikokeishikankyoto (SKK), or placebo for two weeks. This study demonstrated the efficacy of SKK-phytotherapy to reduce PTSD symptoms [41].

We chose Aleozen^®^ in our study for its balanced combination of plant extracts. No study has been published with the product Aleozene which is a certified product in Tunisia. On the other hand, several studies found in the literature have described the effects of each of the components of Aleozene.

*Griffonia simplicifolia* [42, 43], the main component of aleozen, rich in 5 Hydroxytryptophan precursor of serotonin regulating sleep and mood, and *Gentiana lutea* [44-46] with its presence of xanthones is inhibitory of MonoAmine Oxidase, therefore antipsychotic. Rhodiola rosea [47,48], Crataegus oxyacantha [49], Eschscholtzia californica [50] and Melissa officinalis [51] are mild sedatives, Zinc and Vitamin B6 are nutrient for the nerve [52]. The components of aleozene are specific to Aleonat laboratories. In our emergencies, the prescription of this supplement by our physicians has a good feedback.

Our results showed that early administration of Aleozen® reduces occurrence and severity of PTSD symptoms in high risk patients. Also, to make our work original; we initiated treatment in the emergency department. This mirror some previous studies citing trials of Hoge et al who evaluate propranolol, the study of Zohar et al which evaluates hydrocortisone and that of van Zuiden et al. evaluating oxytocin [15, 34, 36].

Several RCTs have demonstrated the effect of herbal medicine in the management of patients who have undergone post-traumatic stress. In the study conducted by Van Zuiden et al, relative to placebo-treated individuals with high acute PTSD symptoms, oxytocin-treated participant had significantly lower PTSD symptom severity. CAPS score decreased from 42.83 in the 10^th^ day to 17.05 after 90 days (reduction was high than 50%) [15]. Second, in a Double-blind, randomised, placebo-controlled study to evaluate the efficacy and safety of a fixed combination containing two plant extracts (Crataegus oxyacantha and Eschscholtzia californica) and magnesium in mild-to-moderate anxiety disorders, Hanus and colleague approved that on whatever parameter (Hamilton total score, Hamilton somatic score, or VAS) the percentage of patients responding to treatment was always significantly greater in the study drug group than in the placebo group (58% vs 43%, p=0.008). [53]

Furthermore, one of the advantages of the study is the homogeneity of the sample with nearly demographic and clinical characteristics in both randomized groups, allowing a valid comparison without selection bias.

In our study, the treatment was administered between the first and the third day after trauma in an arbitrary manner. The optimal time to initiate preventive therapy remains under discussion. Besides, considering the unknown effects of chemoprophylaxis in peoples who would recover naturally after being exposed to a traumatic situation [54], it looks unjustified to provide all patients with preventive treatment. Therefore, targeting only those at high risk of developing PTSD would be a more effective strategy. Indeed, there is promising evidence that quantitative measures can predict the onset of PTSD [55]. Among the strengths of this study is that it had included patients, using PDI, PDEQ and L-CROQ scales, with scores greater than predictive level for PTSD occurrence [19].

### Limitations

There were still several limits in our study; mainly it was a prospective monocentric research. Furthermore, the originality of our study is the herbal medicine treatment that was safe and we did not report any major treatment-related side effects during and after the study. Future multicentric confirmatory studies are needed.

## Conclusion

Through the different results of our study, we have been able to demonstrate that early administration of an herbal supplement prevents the occurrence of PTSD and lowers the severity of symptoms in patients with high risk for development of PTSD.

The results of this work are encouraging and should raise the use of herbal medicine to manage traumatized patient and creating safer health care. Further confirmatory studies are needed.

## Supporting information

consort check list

## Data Availability

All data produced in the present work are contained in the manuscript

## Acknowledgements

All authors thank Aleonat Laboratories, Tunisia.

The authors acknowledge the help of Professor Alexandre Mebazaa in reading this manuscript and his useful comments.

The authors thank all of the physicians who contributed to the study.

