## Supplementary material for "Double-blind, Randomized, Placebo-Controlled study to evaluate the Efficacy of an early treatment with Herbal Supplement based in the Prevention of Post-Traumatic Stress Disorder in emergency department (PHYTéS Study)": suplementary data

| **Table3 : CAPS-5 Items** | | | | |
|---|---|---|---|---|
| | Groups | Moyenne | Ecart-type | P |
| B10days | PLACEBO | 18.8 | 5.1 | 0.991 |
| | ALEOZEN | 18.8 | 5 | |
| C10days | PLACEBO | 7.5 | 2.1 | 0.847 |
| | ALEOZEN | 7.4 | 2.2 | |
| D10days | PLACEBO | 25.2 | 7.4 | 0.745 |
| | ALEOZEN | 24.8 | 7.8 | |
| E10days | PLACEBO | 19 | 7.3 | 0.276 |
| | ALEOZEN | 17.8 | 6.9 | |
| B30 days | PLACEBO | 13.6 | 5.3 | 0.784 |
| | ALEOZEN | 13.3 | 5.6 | |
| C30 days | PLACEBO | 5.3 | 2.3 | 0.999 |
| | ALEOZEN | 5.3 | 2.5 | |
| D30 days | PLACEBO | 17.7 | 7.5 | 0.951 |
| | ALEOZEN | 17.7 | 8.4 | |
| E30 days | PLACEBO | 14 | 5.8 | 0.913 |
| | ALEOZEN | 13.9 | 6.2 | |
| B90 days | PLACEBO | 13.6 | 6.8 | **<0.001** |
| | ALEOZEN | 9.8 | 5.9 | |
| C90 days | PLACEBO | 6.5 | 3.7 | **0.001** |
| | ALEOZEN | 4.7 | 3.1 | |
| D90 days | PLACEBO | 17.5 | 8.8 | **<0.001** |
| | ALEOZEN | 13.1 | 7.1 | |
| E90 days | PLACEBO | 16.8 | 7.2 | **<0.001** |
| | ALEOZEN | 12.9 | 7.3 | |
| B one year | PLACEBO | 12.6 | 6.1 | **0.001** |
| | ALEOZEN | 9.1 | 5.1 | |
| C one year | PLACEBO | 5.6 | 3.7 | **0.041** |
| | ALEOZEN | 2.4 | 2.1 | |
| D one year | PLACEBO | 13.8 | 2.9 | **0.028** |
| | ALEOZEN | 10.7 | 7.6 | |
| E one year | PLACEBO | 12.9 | 8.1 | **0.021** |
| | ALEOZEN | 10.2 | 6.1 | |

| **Table 4 : CAPS-5 Items variations** | | | | |
|---|---|---|---|---|
| | Group | Moyenne | Ecart-type | P |
| BB 30 days | PLACEBO | 5.2 | 5.6 | 0.853 |
| | ALEOZEN | 5.4 | 5.5 | |
| BB 90 days | PLACEBO | 5.6 | 8.8 | **0.006** |
| | ALEOZEN | 9.1 | 6.9 | |
| BB one year | PLACEBO | 6.1 | 4.6 | **0.018** |
| | ALEOZEN | 9.2 | 4.5 | |
| CC 30 days | PLACEBO | 2.1 | 2.2 | 0.768 |
| | ALEOZEN | 2.1 | 2.5 | |
| CC 90 days | PLACEBO | 1.1 | 4.2 | **0.007** |
| | ALEOZEN | 2.7 | 3.5 | |
| CC one year | PLACEBO | 1.9 | 2.6 | **0.038** |
| | ALEOZEN | 2.6 | 3.1 | |
| DD30 days | PLACEBO | 7.5 | 7.8 | 0.693 |
| | ALEOZEN | 7.1 | 8.2 | |
| DD 90 days | PLACEBO | 8.1 | 11.7 | **0.027** |
| | ALEOZEN | 11.7 | 9.3 | |
| DD one year | PLACEBO | 8.8 | 9.1 | **0.044** |
| | ALEOZEN | 10.6 | 8.2 | |
| EE 30 days | PLACEBO | 5.2 | 6.8 | 0.185 |
| | ALEOZEN | 3.9 | 6.3 | |
| EE 90 days | PLACEBO | 2.8 | 9.8 | 0.136 |
| | ALEOZEN | 5 | 9.4 | |
| EE one year | PLACEBO | 2.2 | 6.6 | **0.047** |
| | ALEOZEN | 4.2 | 5.1 | |
